# The UK BiLEVE and Mendelian randomisation: Using multivariable instrumental variables to address “damned if you, dammed if you don’t” adjustment problems

**DOI:** 10.1101/2022.10.25.22281084

**Authors:** Benjamin Woolf, Dipender Gill, Hannah Sallis, Marcus R. Munafò

**Author notes:** These authors contributed equally (Dipender Gill, Hannah Sallis, Marcus R. Munafò). Corresponding Author: Benjamin Woolf, School of Psychological Science, University of Bristol, Bristol, UK.

## Abstract

**Objective:** To explore the use of multivariable instrumental variables to resolve the “dammed if you do, dammed if you don’t” adjustment problem created for Mendelian randomisation (MR) analysis using the smoking or lung function related phenotypes in the UK Biobank (UKB).

**Result:** “dammed if you do, dammed if you don’t” adjustment problems occur when both adjusting and not-adjusting for a variable will induce bias in an analysis. One instance of this occurs because the genotyping chip of UKB participants differed based on lung function/smoking status. In simulations, we show that multivariable instrumental variables analyses can attenuate potential collider bias introduced by adjusting for a proposed covariate, such as the UKB genotyping chip. We then explore the effect of adjusting for genotyping chip in a multivariable MR model exploring the effect of smoking on seven medical outcomes (lung cancer, emphysema, hypertension, stroke, heart diseases, depression, and disabilities). We additionally compare our results to a traditional univariate MR analysis using genome-wide analyses summary statistics which had and had not adjusted for genotyping chip. This analysis implies that the difference in genotyping chip has introduced only a small amount of bias.

## Introduction

Mendelian randomisation (MR) is an increasingly popular method for inferring the causal effect of modifiable exposures on epidemiological outcomes (1). In an MR study, genetic variants which robustly associated with the exposure of interest are used as instruments in an instrumental variables analysis.

One of the most popular resources for conducting MR analyses is the UK Biobank (UKB) (2). The UKB is a large (approximately half a million participants) population cohort study of Britons. The UKB has been used in the MR literature to explore the causal effects of smoking-related phenotypes (3). However, the UKB genotyping was rolled out over several steps.

Participants enrolled in the UK BiLEVE study were genotyped using different instruments (‘genotyping chip’) than other participants (4). Because UK BiLEVE was not randomly sampled, there is a worry that differences in genotyping between participants could cause (confounding) bias if the sampling probability is associated with risk factors for the outcome phenotype of interest. Because of this, there is general advice to adjust UKB GWASs by genotyping chip (5). However, the UK BiLEVE study selected participants who were in a tail or centre of the distribution for lung function and smoking. This poses a problem for genetic designs, like MR, in the UKB exploring smoking and lung function-related phenotypes. If genotyping chip is determined by UK BiLEVE enrolment, which is in turn determined by lung function and smoking status, then adjusting for genotyping chip could introduce collider bias.

Situations where no adjustment will result in bias, but where the required covariate is a collider (see Figure 1) have been described as “dammed if you do, dammed if you don’t” adjustment problems in a recent review of types of covariate controls (6). This review concluded that there were no satisfaction methods for addressing this bias, and they suggested authors should implement sensitivity analyses when encountering this type of problem. Existing guidelines for conducting genetic analysis in the UKB had made similar suggestions of running analyses with and without adjustment for genotyping chip when a phenotype might relate to lung function or smoking (5).

**Figure 1:**
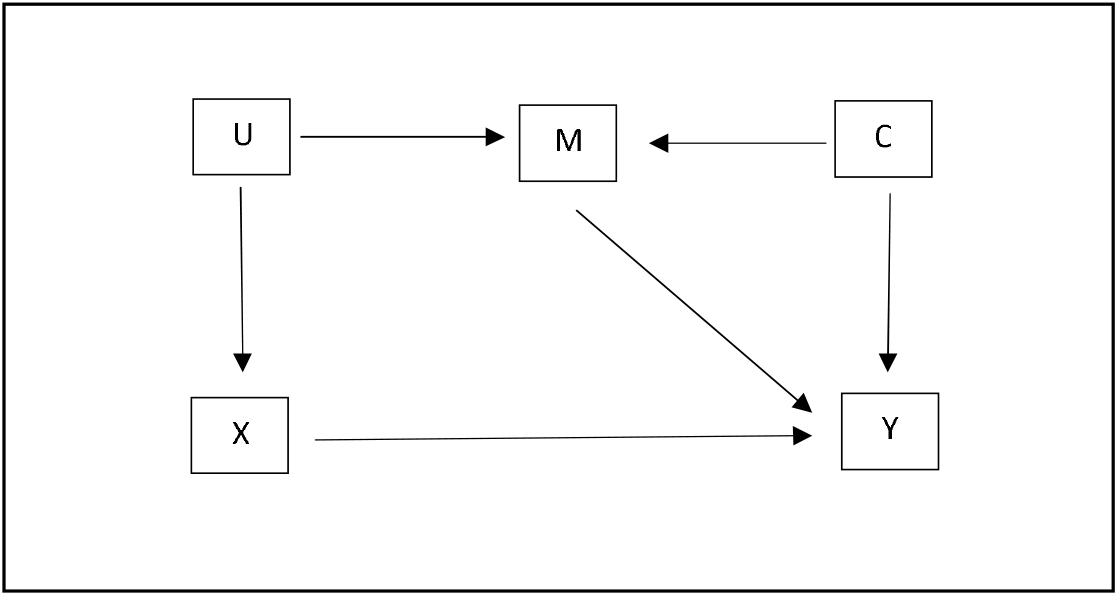
An example “dammed if you do, dammed if you don’t” adjustment problem. Adjusting for M will result in collider bias, but not adjusting for M will introduce confounding.

In this article, we propose a complementary sensitivity analysis to address this type of adjustment paradox, with specific emphasis on addressing bias due to UK BiLEVE in MR studies. Multivariable IV (MVIV) is an extension of traditional IV analysis to include more than one exposure (7). A traditional IV analysis, like MR, assumes that the instrument is robustly associated with the exposure, can affect the outcome only via the exposure, and that there is no ‘back door’ path from the instrument to the outcome. MVIV modifies these assumptions so that: 1) the instrument is robustly associated with the exposure(s) *conditional on the other covariate(s)*, can affect the outcome only via *one of* the exposures, and that, *conditional on all covariates*, there is no ‘back door’ path from the instrument to the outcome. While the effect estimates of a standard IV analysis are the total effect of the exposure on the outcome, MVIV effect estimates should be interpreted as the direct effect of the exposure conditional on the covariates. Because of this, MR applications of MVIV have shown that it can be used to address bias, like collider bias (8), by ensuring that the effect estimate of interest is conditionally independent of a known biasing phenotype. Intuitively then, adding the genotyping chip as a second exposure to an MR model using chip adjusted genome-wide summary statistics should remove any collider bias introduced by adjusting for genotyping chip.

## Main text

### Simulation

#### Aims

We ran a simple simulation to provide a proof of concept that MVIV can be used to address “dammed if you do, dammed if you don’t” adjustment problems. We report our simulations using the ADEMP (aims, data-generating mechanisms, estimands, methods, and performance measures) approach (9).

#### Data-generating mechanisms

We simulated a setting in which there is an exclusion restriction violation (i.e. where the instrument causes the outcome via a path not mediated by the exposure), but were adjusting for this violation would introduce M bias (Figure 2). More formally, we simulated 100 single nucleotide polymorphisms (SNPs, which are common genetic variants) as independent and identically distributed binomial variables with the following parameters:

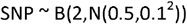

We then simulated two confounders as independent and identically distributed normal variables with the following parameters:

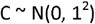

We then defined the exposure as

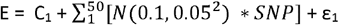

where ε is an error term such that ε_1_ ∼ N(0, 1^2^).

**Figure 2:**
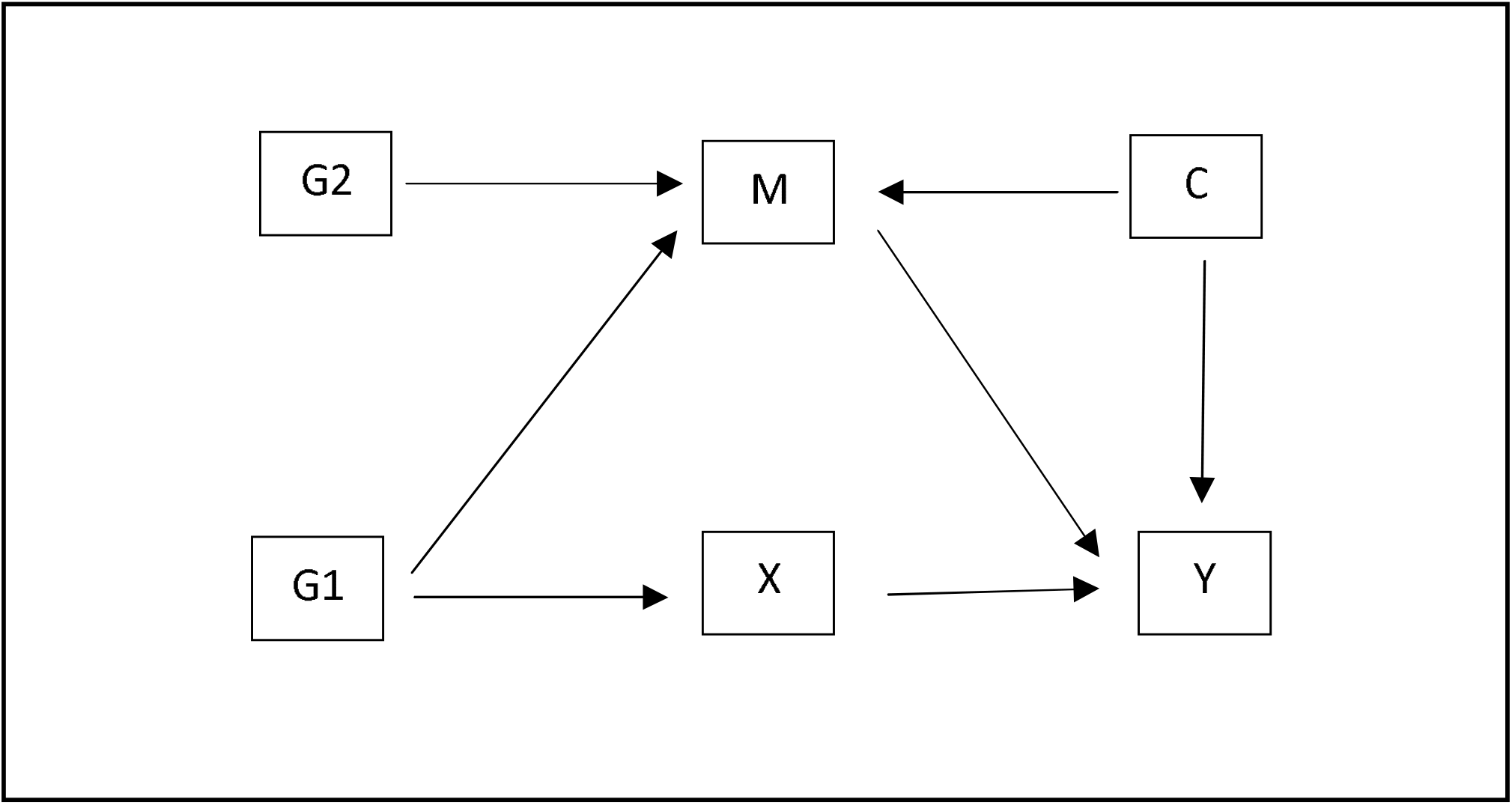
Directed Acyclic Graph of data generative model used in simulation. Here, G1 is the genetic liability to the exposure, and G2 the genetic liability to the potential covariate. C is the potential covariate, and U1 and U2 are confounders. X and Y are the exposure and outcome respectively. M is additionally pleotropic and a collider.

We defined the potential covariate for blocking the exclusion restriction violation as

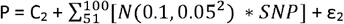

where ε_2_ ∼ N(0, 1^2^).

Finally we defined the outcome as

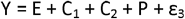

where ε_3_ ∼ N(0, 1^2^).

#### Estimands

The causal effect of the exposure on the outcome.

#### Methods

We compare three methods for estimating the causal effect of the exposure on the outcome:

1. We ran an inverse variance weighted (intercept free) regression of B_y_ ∼ B_x_ + 0, where B_y_ is the SNP-outcome association and B_x_ is the SNP-exposure association, and where the B_x_ and B_y_ were estimated in linear models which additionally adjusted for P.
2. An inverse-variance weighted (intercept free) regression of B_y_ ∼ B_x_ + 0, where the B_x_ and B_y_ were not estimated in linear models which did not additionally adjust for P.
3. An inverse-variance weighted (intercept free) regression of B_y_ ∼ B_x_ + B_p_ + 0, where B_p_ is the SNP-covariate association, and where the B_x_ and B_y_ were estimated in linear models which additionally adjusted for P.

For readers less familiar with the MR literature, it is worth noting that the intercept-free weighted regression is equivalent to an inverse variance weighted meta-analysis of the IV Wald ratios for each SNP. In addition, (1) and (2) only used the first 50 simulated SNPs (i.e. those which associated with the exposure), while (3) used all 100 simulated SNPs (i.e. SNPs which associated with either exposure). B_y_, B_x_, and B_p_ were all estimated in non-overlapping samples of 250,000 participants.

#### Performance measure

The mean bias in the causal effect of the exposure on the outcome over 1000 iterations.

#### Results of the simulation

The simulation found that both adjusting and not adjusting the linear model for the covariate resulted in bias (mean bias = -0.445 [MC SE = 0.002] and 0.972 [MC SE = 0.001] for the adjusted and not adjusted analysis respectively). On the other hand, the MVIV model attenuated most of the bias (mean bias = -0.064 [MC SE = 0.002]).

### Applied example with the UKB genotyping chip

We used a two-sample MR analysis of the effect of smoking on seven outcomes (lung cancer, emphysema, depression, hypertension, stroke, heart diseases, and diabetes) in the UKB as an applied example. The outcomes were chosen because there is existing literature implying a causal association between smoking and these outcomes (10–15). We ran three versions of this analysis: 1) using univariable MR to estimate the effect of smoking on the outcomes where the UKB smoking, lung cancer and emphysema GWASs had adjusted for genotyping chip, 2) using univariable MR to estimate the effect of smoking on the outcomes where the UKB smoking, lung cancer and emphysema GWASs had not adjusted for genotyping chip, and 3) using multivariable MR estimate the effect smoking on the outcomes adjusted for genotyping chip where the UKB smoking, lung cancer and emphysema GWASs had adjusted for genotyping chip.

We used the Wootton et al UKB lifetime smoking GWAS as a source of SNP-exposure associations (3), which we by dividing the effect estimate and standard error by 0.6940093. To estimate genotype-chip association we ran a GWAS (adjusted for age, sex, and the first 10 principal components of ancestry) using BOLT-LMM in the MRC-IEU UKB GWAS pipeline. Full methods for both GWASs described elsewhere (3,16). In both the univariate and multivariate setting, we selected genome-wide significant SNPs associated with the exposure(s) of interest as genetic instruments, and then clumped this list using an r^2^ of 0.001 and kb of 10,000. We additionally implemented the FIQT WCC on the exposure GWASs to correct for any effect of Winner’s curse (17).

We used the Elsworth’s UKB GWASs in the MRC-IEU OpenGWAS platform as a source of SNP-emphysema and -hypertension associations (18); as well as Nikpay et al’s GWAS of CAD, Malik et al’s GWAS of stroke, Wang et al’s GWAS of lung cancer, Howard et al’s GWAS of depression and the FinnGen round 5 GWAS of disabilities (19–23). Details on genotyping, quality control, and phenotyping can be found in the original publications and on the UKB website (https://biobank.ndph.ox.ac.uk/ukb/search.cgi). All outcome GWASs were on the odds ratio scale. We harmonised the exposure and outcome samples, and removed palindromic SNPs whose effect allele could not be inferred using based on minor allele frequency. We used four MR estimators: IVW, weighted mode, weighted median, and MR Egger. We additionally estimated the heterogeneity in the MR Wald ratios using the Cochrane Q statistic as a control for exclusion restriction violations. The univariate MR analysis was implemented using the TwoSampleMR R package (24,25). Multivariable MR analyse additionally used the MVMR, MendelianRandomsiaiton, and MVMRMode R packages (26–28).

Table 1 presents the results of this analysis. These broadly show highly consistent findings across the three methods, with most of the changes in estimates smaller than the standard error of each point estimate. Overall, this would therefore broadly imply that there is minimal collider or confounding bias introduced by adjusting or not adjusting for genotyping chip.

**Table 1:**
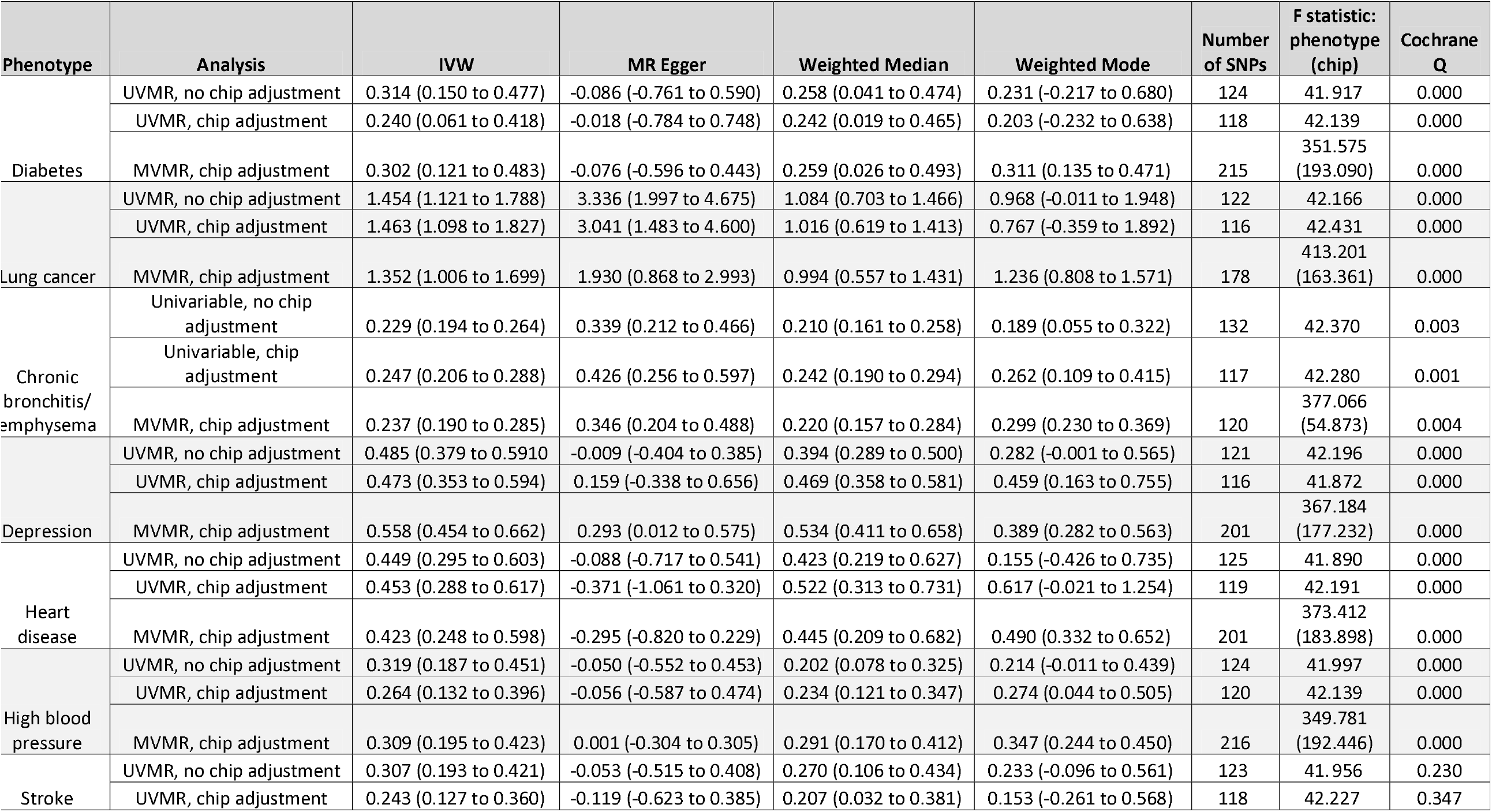
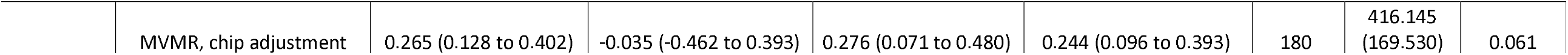
Results of applied example. UVMR = univariable Mendelian randomisation, MVMR = multivariable Mendelian randomisation.

### Limitations

Here we have shown that MVIV can, in theory, be used to attenuate bias when “dammed if you do, dammed if you don’t” adjustment problems occur in an IV analysis. We then apply this to the UKB and show that, despite differences in genotyping depending on lung function and smoking status of participants, the UK BiLEVE study appears to have introduced only a small amount of bias into our estimates of the causal effect of smoking on lung cancer.

There are three complications to the application of our findings to address differences in genotyping chip in the UKB, which we believe mean that our proposal should be used to supplement, rather than replace the existing guidance of performing both a chip-adjusted, and no-chip-adjusted, analysis as a sensitivity analysis. Firstly, MVIV has additional parameters than univariable IV analyses, and will therefore be even less precise. Secondly, the collinearity of exposures (such as smoking and genotype chip) can also introduce conditionally weak instruments into analyses which would have strong instruments in a university setting. Although not an issue in our applied analysis, this could become a major issue in analysis using weaker instruments, such as parental smoking status. Hence, authors should come to a judgment about which method will have a lower mean squared error and then use the alternatives as sensitivity analyses. Thirdly, because enrolment into the UK BiLEVE study was determined by smoking and lung function, it could be argued that it is, in effect, a proxy of these variables. If this is the case, then adjusting for genotyping chip in a model would potentially do something equivalent to adjusting for a mediator in a traditional regression analysis, and therefore introduce bias. This underpins the importance of not using the MVIV analysis to replace the existing guidelines.

A final, but related, limitation when applying our proposal to other settings is that there has to be a way to validity instrument the proposed covariate. Since there are many settings, especially when using summary data IV analysis like two-sample MR, when study-specific variables, such as UKB genotyping hip, this may be more common than the authors would hope.

## Data Availability

The code and GWAS summary statistics used in this study is available from https://doi.org/10.17605/OSF.IO/RPBYD. The GWAS summary statistics will also be made available from the MRC-IEU OpenGWAS project.
This project was conducted using UK Biobank application no. 15825. UK Biobank was established by the Wellcome Trust medical charity, Medical Research Council, Department of Health, Scottish Government and the Northwest Regional Development Agency. It has also had funding from the Welsh Government, British Heart Foundation, Cancer Research UK and Diabetes UK. UK Biobank is supported by the National Health Service (NHS). UK Biobank is open to bona fide researchers anywhere in the world.

## Declarations

### Ethics approval and consent to participate

UKB received ethical approval from the North West Multi-Centre Research Ethics Committee (REC reference 11/NW/0382). All participants provided written informed consent to participate in the study. Data from the UKB are fully anonymised.

### Consent for publication

All authors consent to this work being published.

### Availability of data and materials

The code and GWAS summary statistics used in this study is available from https://doi.org/10.17605/OSF.IO/RPBYD. The GWAS summary statistics will also be made available from the MRC-IEU OpenGWAS project.

This project was conducted using UK Biobank application no. 15825. UK Biobank was established by the Wellcome Trust medical charity, Medical Research Council, Department of Health, Scottish Government and the Northwest Regional Development Agency. It has also had funding from the Welsh Government, British Heart Foundation, Cancer Research UK and Diabetes UK. UK Biobank is supported by the National Health Service (NHS). UK Biobank is open to bona fide researchers anywhere in the world.

### Authors’ contributions

BW conceived of and designed the study. All authors contributed to the writing of the manuscript.

## Acknowledgements

This work was carried out using the computational facilities of the Advanced Computing Research Centre, University of Bristol -http://www.bris.ac.uk/acrc/.

We are very grateful to Kate Tilling for providing feedback on the simulation methods and interpretation of the study.

